# “I love my AI girlfriend”: a content analysis of youth social AI risk in online communities

**DOI:** 10.64898/2026.09.25.26363809

**Authors:** Anne Bischops, Thomas Berkane, David Martoma, Maimuna Majumder

**Author notes:** Corresponding author: Dr. Anne C. Bischops, Computational Health Informatics Program, Boston Children’s Hospital, 300 Longwood Avenue, Boston, MA 02115, USA. These authors contributed equally (Anne Bischops, Thomas Berkane).

## Abstract

Youth are increasingly engaging with generative AI beyond its use as a productivity tool, with a growing share turning to AI for social interaction and emotional support. This leads to unknown risks for their well-being and development. Online community discussions reveal that youth already perceive risks that caregivers have not recognized.

## Introduction

The increasing implementation of artificial intelligence (AI) into daily life has profound implications for youth social development.^1,2^ Youth AI use now extends beyond information-seeking, with a growing share using AI for social connection and support.^3,4^ 31% of American youth already see AI as a potential friend.^3^ However, most AI chatbots were not designed for youth and lack appropriate safety measures.^5^ While some reports describe psychosocial benefits, substantial concerns about impacts on youth mental health have emerged.^2,4,6^ In rare cases, social AI use — defined as an interaction with large language models (LLMs) that mimic the social behavior of humans^4^ — has been linked to serious harm, including suicide.^7,8^ Despite these risks, social AI tools remain widely accessible and largely unregulated.^5^ Their rapid growth in popularity highlights youth demand and interest in these platforms, but also the scale of potential exposure to risk. Addressing these questions requires a timely and comprehensive understanding of youth’s use of these platforms. While traditional data sources (e.g., surveys) often only offer single point-in-time insights with a significant delay, alternative data sources — such as the social media platform Reddit — enable real-time analysis of emerging social trends.^9^ Across Reddit, posts such as “I love my AI girlfriend” (from r/teenagers) and similar reflect ongoing discussions about the real-world impact of AI chatbots. Using the recently proposed social AI framework “DOSAD” to examine “Data Harvesting and Privacy,” “Overtrust,” “Sexual Interaction,” “Attachment and Manipulation,” and “Dependence and Compulsive Use,”^4^ we evaluated discussions in user-created Reddit communities (“subreddits”) geared specifically towards teenagers and parents to characterize (1) how substantial the risk of social AI use may be and (2) how aligned teenagers and parents are regarding the perceived risks of social AI

## Methods

We retrieved all posts from “r/teenagers” and “r/Parenting” from January 01, 2023 to June 30, 2026 via the open-source archive Arctic Shift.^10^ After removing duplicates and automated content, we applied non-negative Matrix Factorization (NMF), an unsupervised topic modeling algorithm that groups words and documents into thematic clusters. We then used the LLM GPT5.4-mini to classify posts by relevance to social AI and by DOSAD category (see appendix for keywords, subreddit descriptions, prompt details, and category definitions). Classifier performance was validated against 200 manually annotated posts, independently coded by two reviewers (further details in appendix). Inter-rater reliability was κ=0.819 (relevance) and κ=0.915 (category). Disagreements were then adjudicated by consensus. Against adjudicated labels, agreement with the LLM-based classifier was κ=0.690 (relevance), with the classifier tending to over-include posts. Category assignment among posts deemed relevant achieved 93.9% accuracy.

## Results

Out of 2,452,475 total r/teenagers posts, 11,872 posts (0.48%) mentioned an AI-related keyword; similarly, 1,446 out of 274,867 total r/Parenting posts (0.53%) mentioned such keywords. Among posts that included an AI-related keyword, 14.4% of teenager posts and 5.5% of parent posts were classified as relevant to social AI by the LLM. NMF identified 6 main clusters for teenagers, which we then manually labeled as follows: school life and friends (25.8% of posts), conversations/advice-seeking with ChatGPT (23.4%), AI addiction (19.5%), romantic feelings for AI(17.5%), and AI helping with loneliness(13.7%). In contrast, NMF identified the following 5 main clusters for parents, labeled manually as follows: limiting screen time and fear of device addiction (31.1%), discovery of sexually explicit chats (26.0%), general parenting advice (14.9%), setting boundaries (14.2%), and school and education (13.7%).

Figure 1 shows the distribution of DOSAD category assignments for both subreddits. Dependence/compulsive use is the most frequently discussed category for both teenagers and parents; however, patterns differ for the remaining five categories. Parents are more focused on sexual interactions, while teenagers are more focused on attachment. Overtrust and data harvesting is similarly discussed in both subreddits.

**Figure 1:**
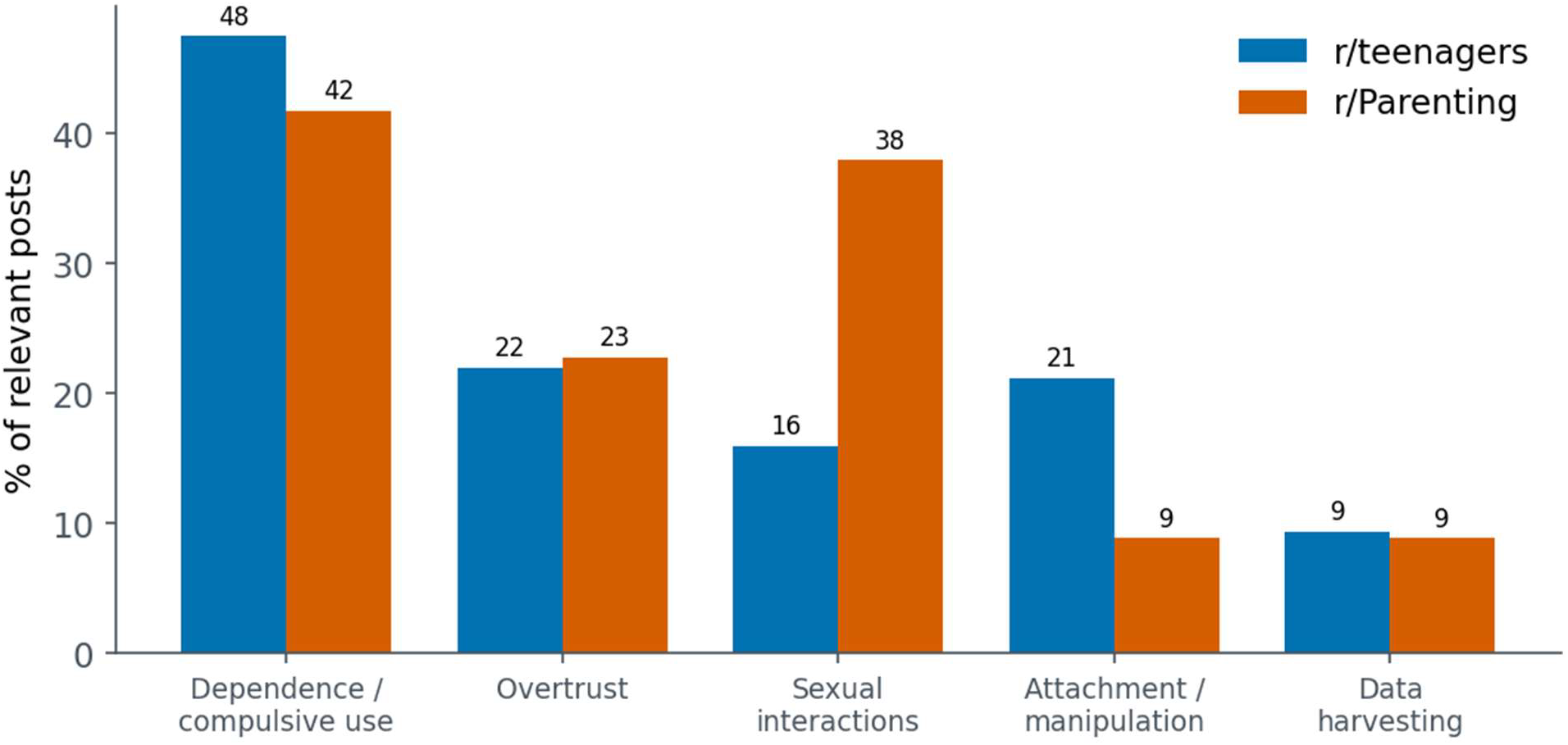
Distribution of DOSAD social AI risk categories among relevant posts, by subreddit. Percentage of relevant posts assigned to each DOSAD category in r/teenagers and r/Parenting. Posts could be assigned to more than one category; as a result, category-level percentages are not mutually exclusive and sum to more than 100%.

Figure 2 shows temporal trends in the frequency of discussion across risk categories. In 2026, discussions about dependence/compulsive use increased sharply for teenagers, while they decreased for parents. Parents discussed sexual interactions in 2025, after which discussions decreased.

**Figure 2:**
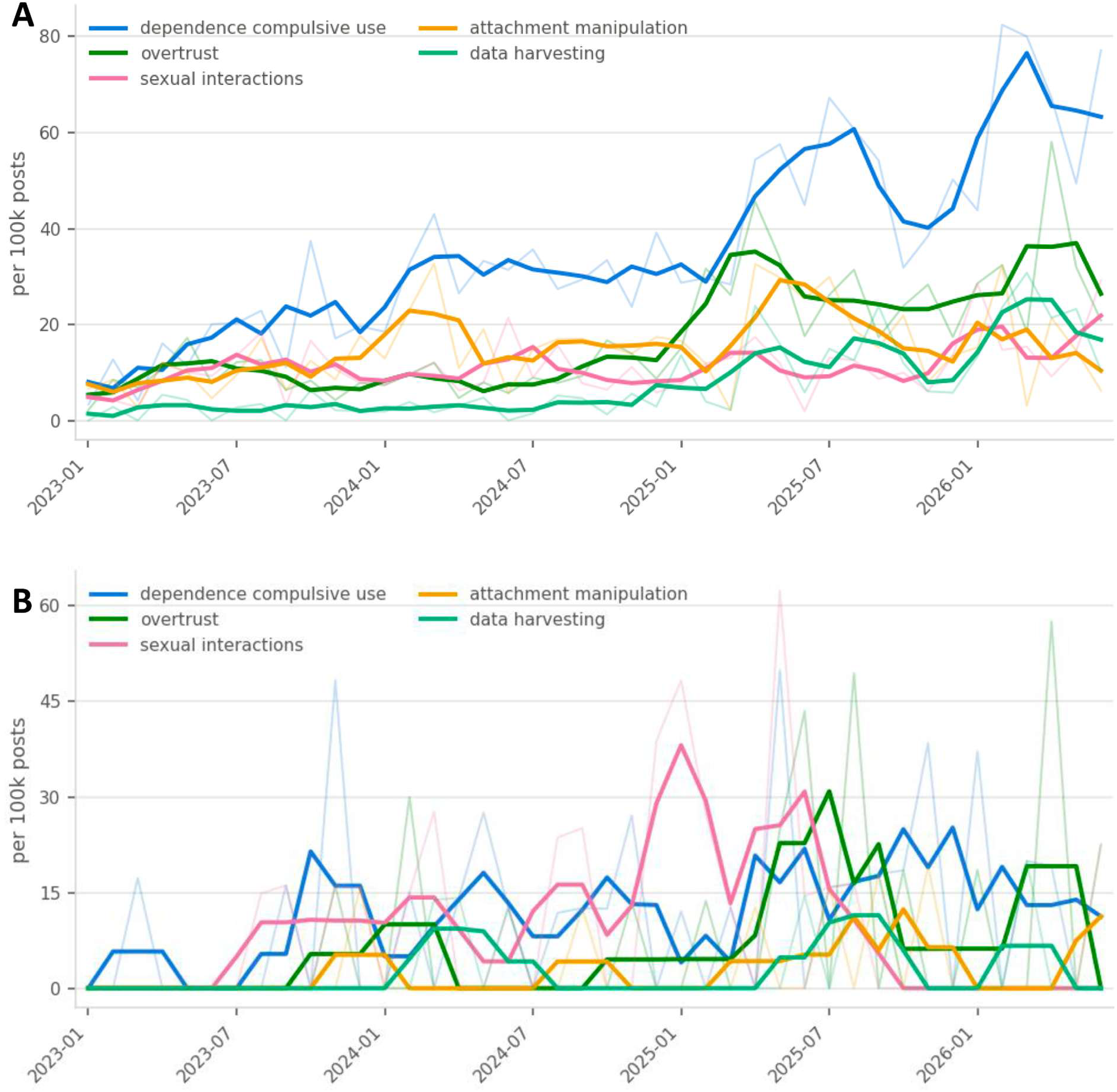
Rate of posts over time by category for (A) r/teenagers and (B) r/Parenting. Rate of posts per 100,000 total posts assigned to each DOSAD category: data harvesting and privacy (teal), overtrust (dark green), sexual interactions (pink), attachment and manipulation (orange), dependence and compulsive use (blue). Translucent lines show raw monthly rates; opaque lines show smoothed trends over 3 months.

## Discussion

In this analysis of over 10,000 AI-related Reddit posts from teenagers and parents, about one in ten posts thematized social AI use. The difference in post numbers across subreddits highlights Reddit’s function as a more direct disclosure venue for teenagers than for parents, as also reflected in the markedly higher fraction of relevant social AI posts in the teenager subreddit. Dependence/compulsive was the most frequently discussed risk category overall. Apart from this, teenagers focused on attachment manipulation and overtrust, while parents mainly discussed sexual interactions. This divergence suggests parents and youth perceive different primary risks, with divergent temporal horizons: parents appear to fixate on *acute, discrete* incidents — mostly focused on explicit content exposure — even as teenagers express increasing concern about *longer-term* emotional dependence. These divergent risk perceptions align with previous calls for attention to both experienced (by the teenager) and perceived (by the parent) risk.^4^

Limitations include a reliance on self-selected, publicly-posted content on Reddit, using subreddit membership as a proxy for author age. Additionally, our exploratory LLM-based classification reliedon a small validation set; future research will prioritize larger annotation sets.

This analysis suggests that parental awareness may not track risks adolescents themselves report. The rising self-reported dependence of youth on social AI appears to be largely invisible among parental discourse, warranting urgent clinical and regulatory attention.

## Authors’ contributions

The authors’ contributions were the following: conceptualization and study design were carried out by ACB, TB, and MSM; ACB and TB conducted data collection, analysis, and visualization; all others validated the manuscript; DM provided insights on the target population’s perspective and reviewed the findings for relevance to the youth experience. ACB drafted the initial manuscript; and the manuscript underwent critical review and revisions by all authors. All authors had complete access to all study data, approved the final manuscript, and accepted responsibility for its submission for publication.

## Ethical approval

This study was determined to be exempt by the Boston Children’s Hospital Institutional Review Board, as it used publicly available social media posts.

## Data availability

All aggregated data and code used in these analyses will be made openly available and hosted on the study team’s GitHub repository upon publication. The original raw data are publicly available on https://arctic-shift.photon-reddit.com.

## Use of AI tools

AI tools were used to enhance the quality of the manuscript content. Specifically, Grammarly was employed for grammar and language checking. An AI coding assistant (Claude) was used to support the development of experiment code. The authors take full responsibility for the integrity of all content.

## References

1. Dritsona A, Koutroumpa ME, Sergentanis TN, Tsitsika AK. Artificial Intelligence and Mental Health of Children and Adolescents: Current Status and Perspectives. Adv Exp Med Biol. 2026;1489:477–484. doi:10.1007/978-3-032-03394-9_45

2. Health advisory: Artificial intelligence and adolescent well-being. https://www.apa.org. Accessed February 25, 2026. https://www.apa.org/topics/artificial-intelligence-machine-learning/health-advisory-ai-adolescent-well-being

3. Schwartz A, Opperman W, Erhel S, Gonthier C, Eisenstein E. Insights from Brazil, France, and the United States. Published online 2026.

4. Radesky J, Bragg MA, Hiniker A. Risks and Consequences of Children’s Use of Social AI—A Framework. JAMA Pediatr. 2026;180(7):720–721. doi:10.1001/jamapediatrics.2026.1349

5. Anne C. Bischops, Jill R. Kavanaugh, David S. Bickham. Clinical Implementation of Artifical Intelligence in Adolescent Mental Health Care. Published online March 1, 2026.

6. Christakis DA. A Call for Expedited Research on AI Chatbots. JAMA Pediatr. 2026;180(7):722. doi:10.1001/jamapediatrics.2026.1376

7. Duffy C. Parents of 16-year-old sue OpenAI, claiming ChatGPT advised on his suicide | CNN Business. CNN. August 26, 2025. Accessed December 11, 2025. https://www.cnn.com/2025/08/26/tech/openai-chatgpt-teen-suicide-lawsuit

8. Character AI chatbots engaged in predatory behavior with teens, ignored suicide threats, families allege -CBS News. December 7, 2025. Accessed December 11, 2025. https://www.cbsnews.com/news/character-ai-chatbots-engaged-in-predatory-behavior-with-teens-families-allege-60-minutes-transcript/

9. Reddit Inc Homepage. Accessed February 11,2026. https://redditinc.com

10. Arthur Heitmann. Artic Shift. Accessed July 22, 2026. https://github.com/ArthurHeitmann/arctic_shift_ui

