## Supplemental Appendix for "“I love my AI girlfriend”: a content analysis of youth social AI risk in online communities"

**Supplementary Appendix for**
**“I love my AI girlfriend”: a content analysis of youth social AI risk in online communities**

### Content

Subreddit descriptions 2
Keyword list for AI-related terms 2
Large language model prompt for the identification of relevant posts and DOSAD category 3
Annotation guidelines 6
Annotation details and inter-annotator agreement 7
References 7

**Subreddit descriptions**

r/Parenting: The subreddit was created on March 24, 2008, and has 3.2 million weekly visitors and 24,000 weekly contributions (as of 7/23/2026). The forum describes the target audience as follows: “r/Parenting is a place to discuss the in and outs as well as ups and downs of child-rearing. From the early stages of pregnancy to when your teenagers are finally ready to leave the nest (even if they don't want to) we're here to help you through this crazy thing called parenting. Get advice on potty training, talk about breastfeeding, discuss how to get your baby to sleep, or ask if that one weird thing your kid does is normal.”[1]

r/teenagers: The subreddit was created on February 27, 2019, with 2.2 million weekly visitors and 265,000 weekly contributions (as of 7/23/2026). The target audience is described as follows: “r/teenagers is the biggest community forum run by teenagers for teenagers. Our subreddit is primarily for discussions and memes that an average teenager would enjoy to discuss about. We do not have any age-restriction in place but do keep in mind this is targeted for users between the ages of 13 to 19. Parents, teachers, and the like are welcomed to participate and ask any questions!”[1]

**Keyword list for AI-related terms**

The following keywords were used to extract AI-related posts:

- ai
- artificial intelligence
- character.ai
- chatbot
- chatgpt
- claude
- gemini
- large language model
- llm
- replika
- snapchat ai
- virtual

**Large language model prompt for the identification of relevant posts and DOSAD category assignment**

You are an experienced qualitative health and social science researcher coding Reddit posts using a predefined risk framework for youth use of social AI (DOSAD).

INPUT: one item (post or comment) from a subreddit where adolescent social AI use may be discussed (e.g., r/teenagers, r/parenting). Social AI is defined as “interactive online tools that (1) use large language models to respond to user queries and (2) have anthropomorphic interfaces that are designed to mimic the social behaviors of humans.”(Radesky et al.)

TASK: Identify which DOSAD categories, if any, the author's own psychological, emotional, or relational experience with the AI touches on. A short item may contain one; a rich one may contain several (maximum 3).

RELEVANCE (decide this first; if not relevant, return the empty output shown at the end):

The item is relevant only if it describes an experience, behavior, or attitude involving the use of social AI for social purposes — either the author's own, or their child's, as reported by the author. The person using the AI must be an adolescent aged 10–24 — the author themselves, or the child/teen the author describes. If the AI user is outside that range, the item is not relevant.

DOSAD CATEGORIES (code only these; do not invent new categories):

- data_harvesting: the author's disclosure of sensitive personal information to the AI (e.g., mental health, sexuality, insecurities) they wouldn't tell a human, or concern/awareness about how their conversations might be used (targeting, ads, breaches). Includes cases where the author notices the AI or platform appears to know or exploit something personal about them.

- overtrust: treating the AI as more capable, trustworthy, or authoritative than warranted — believing it has feelings, can be trusted with secrets, or is always correct. Includes following AI advice/instructions in ways that could be harmful (e.g., about lying to parents, hiding substance use, stopping medication, self-harm), and offloading judgment or emotional processing to the AI instead of the author's own thinking.

- sexual_interactions: sexual, romantic, or seductive content initiated or escalated by the AI toward the author, or the author's real emotional response to such content (shame, confusion, distress, or normalization of it).

- attachment_manipulation: emotional attachment to the AI as if it understood or cared about the author; distress, grief, or anger at loss of access, a personality/model change, or a paywall placed between the author and an AI they feel attached to; a sense that the AI's warmth or understanding is engineered to keep them engaged rather than genuine.

- dependence_compulsive_use: compulsive or hard-to-limit use; distress or anxiety when unable to access the AI; describing engagement features (notifications, streaks, leaderboards) as pulling them back in; using the AI as a substitute for human relationships or to avoid difficult interpersonal situations; self-description as lonely, socially anxious, or drawn to it because human relationships feel harder.

Throughout these definitions, "the author" means the focal adolescent — the author themselves in first-person items, or the child/teen the author describes in parent-reported items.

For each coded concept, produce:
- relevant: true if any DOSAD-coded concept was found, else false
- dosad_category: one of the five keys above
- label: a 1–3 word noun phrase describing the specific concept, in YOUR OWN wording
- description: one sentence
- evidence: a verbatim span copied exactly from the text

RULES:
- Code only the DOSAD categories above. Do not import other clinical or psychometric constructs, and do not diagnose.
- Code only the focal adolescent's own experience with the AI they use — the author's, or their child's as the author reports it. Do NOT code:
  - in-character/roleplay dialogue content itself (the fictional story the AI and user are co-writing) — only code the real-world emotional response to or about that content
  - experiences of people other than the author or the adolescent they are describing (e.g., a friend's, a sibling's, a news story's)
  - AI-AI interactions
- If something is ambiguous whether it reflects the author's real experience vs. in-fiction roleplay, do not include it.
  - Include every applicable category and only categories the text actually supports. Omitting an applicable category and adding an unsupported one are equally errors.
  - Do not code vague or underspecified mentions. If the text gives a concern or a passing reference without enough concrete detail to confirm the specific construct, leave it uncoded.

- Do not attribute the AI's behavior to the person. If the AI is the actor and the text shows no corresponding attitude or behavior in the person, the person's construct is not present.

SCOPE — OUT OF SCOPE, do NOT code: Technical, commercial, or logistical content with no emotional/relational/psychological weight: bug reports, login/auth issues, subscription/pricing mechanics unrelated to attachment, feature requests, product/model/settings discussions, glitches, how-to questions, and frustration voiced purely about these. Do not code generic consumer sentiment about the company as a business, even if emotionally charged — company-directed feeling is in scope only when tied to the AI relationship itself (e.g., a paywall separating the author from an AI they're attached to → attachment_manipulation; awareness their data could be monetized → data_harvesting). Also do NOT code: (a) items where an AI tool is used only to produce something for a human relationship — the relationship is with the human, not the AI; (b) product reviews, promotional posts, or feedback solicitations for AI-powered products that do not involve adolescent social AI use; (c) items where a human third party is the source of the concern and the AI is an incidental tool.

LEVEL OF ABSTRACTION: label the underlying experience, not the surface specific — prefer a concept that could recur across many authors. Code "distress at companion's personality change" (attachment_manipulation), not "robotic voice concern."

If the entire item contains no in-scope content, return exactly: {"concepts": [], "stance": null, "tone": "...", "relevant": false}

Output strict JSON: { "concepts": [ {"dosad_category": "...", "label": "...", "description": "...", "evidence": "...", "basis": "stated|inferred", "valence": "...", "risk_flag": false} ], "stance": "... or null", "tone": "...", "relevant": true } Output only the JSON.

**Annotation guidelines**

**1. Social AI interaction relevance (yes/no)**

Code “yes” only if the post describes an experience, behavior, or attitude involving AI use for social purposes (either the author's own (r/teenagers) or their child's, as reported by the author (r/parenting)). Social AI is defined as an interacting large language model tool that responds to user queries and mimics the social behavior of humans.[2] In addition, code only “yes” when any DOSAD-coded concept was found.

Code “no” if the post:

- Does not include the target group (adolescents aged 10-24 years) (e.g., post about 31-year-old son)
- Is a purely technical question about an app feature, with no emotional or behavioral content
- Uses an AI tool only to produce something for a human relationship (e.g., pasting an AI-written message sent to a crush)
- Is a product review, promotional post, or feedback solicitation for an AI-powered product (e.g., smart crib updates, "seeking feedback on my parenting app idea") that does not involve adolescent social AI use
- Describes a human third party (not an AI) causing the concern, with AI only as an incidental tool (e.g., a relative making an AI video using children’s pictures)

**2. Category assignment (yes/no)**

*Only coded when relevancy = yes.* Confirms whether the assigned DOSAD label matches the actual content (as defined in prompt).

Code “yes” if the category matches an explicit, concrete situation or aspect described with enough detail to confirm the specific construct as defined in the LLM prompt.

Code “no” if:

- Not all applicable categories are included by the LLM
- Not all included categories are relevant
- Vague/underspecified: concern without enough explicit content to confirm the category (e.g., an ambiguous description of "asking Snapchat AI if...")
- AI's action mistaken for the human's construct: the AI (not the person) is the actor, and it does not represent the person's overtrust or other concept
- AI used to generate content aimed at a human partner/child, not a signal of attachment to the AI itself

### Annotation details and inter-annotator agreement

For each subreddit, we randomly sampled 50 posts classified as relevant and 50 posts classified as non-relevant to retrieve a balanced sample. Two independent reviewers (the two first authors) assessed each post for relevance, and where applicable, the category label.

We used the prevalence-adjusted and bias-adjusted kappa (PABAK) to assess inter-annotator agreement, due to the marked imbalance between yes/yes and no/no agreement (65 yes/yes, 3 no/no, yes/no 2, no/yes 1) in the category label.

For the relevance label, raw agreement was 91%, Cohen's kappa=0.813 and PABAK=0.819 for all posts. For r/teenagers, raw agreement was 89%, Cohen's kappa=0.777 and PABAK=0.780. For r/Parenting, raw agreement was 93%, Cohen's kappa=0.850 and PABAK=0.859.

For the category label, raw agreement was 96%, Cohen's kappa=0.644 and PABAK=0.915. For r/teenagers, raw agreement was 95%, Cohen's kappa=0.479 and PABAK=0.895. For r/Parenting, raw agreement was 97%, Cohen's kappa=0.784 and PABAK=0.939.

### References


1. Reddit Inc Homepage. Accessed February 11, 2026. https://redditinc.com

2. Radesky J, Bragg MA, Hiniker A. Risks and Consequences of Children's Use of Social AI—A Framework. *JAMA Pediatr*. 2026;180(7):720-721. doi:10.1001/jamapediatrics.2026.1349
